# Real-world comparison of counterfactual inference methods for evaluating impact of antidepressants on COVID outcomes

**DOI:** 10.1101/2025.10.13.25337938

**Authors:** Sonja Kleper, Mamoon Habib, Anugrah Vaishnav, Tingjian Ge, Rachel Melamed

## Abstract

In order to discover drugs that could be repurposed for a public health emergency like the COVID-19 pandemic, the National COVID Cohort Collaborative (N3C) database compiles health records including 22 millions individuals and 8.9 million cases of COVID-19. Here, we sought to use this data to systematically investigate whether antidepressants could impact COVID-19 outcomes, adjusting for known risk factors for severe outcome. We conducted large scale target trial emulation, comparing all pairs of 18 antidepressants to each other. Because the best approach for discovering such drug effects from observational data is not known, we applied a series of methods for identifying drug effects by estimating the counterfactual outcome that would be observed in a randomized trial. We found that all methods for counterfactual outcome estimation were prone to bias due to poorly controlled unmeasured confounding. We describe this bias, which appears to be induced partly by conditioning on treatment exposure, opening a back door path, via collider effects, between treatment and exposure via unmeasured confounders. Via empirical simulations, we show that our approach is able to detect this bias. In result, we state that we are not able to confidently identify any antidepressant impacting COVID-19 outcomes.

## Introduction

At the start of the COVID-19 pandemic, there was a lack of effective treatments, and high rates of severe outcomes^1^. In order to rapidly advance management of the disease, studies aimed to assess if antihypertensives^2^, antidepressants^3^, or, controversially, hydroxychoroquine^4^ could be beneficial for COVID-19. To enable observational data analysis, the National COVID Cohort Collaborative (N3C), created a centralized database of health records including 22 millions individuals and 8.9 million cases of COVID-19. A number of studies have mined this database to assess if commonly used drugs could change COVID-19 outcome.^5,5–8^ This intensive interest in rapid data-driven discovery of promising therapies aimed to overcome the weaknesses of clinical trials: trials are slow and expensive, making them poorly suited for uncovering unexpected drug effects, particularly in a rapidly developing public health crisis. However, it is widely recognized that clinical trials are needed to rigorously test a drug’s efficacy in humans. Crucially, clinical trials include randomized assignment of treatment, which removes any confounding association between health history and drug prescription, allowing identification of the true effect of a drug. To discover such drug effects using observational health data, *causal inference* methods aim to emulate the result that a randomized clinical trial would have obtained, if it had been conducted. However, these methods have not been deployed and compared systematically in order to understand their strengths and weaknesses for discovering hidden effects of drugs on health outcomes.

Here, we compare multiple methods for estimating the *counterfactual outcome*, defined as the outcome that a patient would have had, if they had been exposed to a drug. These methods typically model a patient’s health history in order to forecast the disease outcome that would have occurred after drug exposure. If the model is successful, then, conditional on patient history, there should be no association between which treatment the individual received and their predicted outcome. Then, this method is able to remove the confounding bias and recover the true drug effect. Many such models exist, including “metalearners”, which describes model designs that can encompass a range of models. Recent methods apply deep learning to improve these estimates. We propose that by deploying these methods for identifying drugs with promising effects on outcomes for COVID-19, we can better understand their performance and potential for drug discovery. We compare traditional regression, machine learning, and deep learning methods for counterfactual outcome prediction. To this end, we propose that simultaneously estimating a range of drug effects will provide a basis for this comparison. We focus on antidepressants as they are a widely used class of medications that have been previously investigated for possible benefit for COVID-19. Therefore our contributions are 1) conducting large-scale emulation of target trials to obtain effect estimates from a range of models for counterfactual effect estimation; 2) developing a framework for assessing the models using the results of the target trial emulation; 3) integrating results across multiple models to test for robustly supported drug associations with COVID-19 outcome.

## Methods

### Preparation of data for active comparator target trial emulation

We integrated the patient records with the analysis tables available from the study by Bennett et. al.^1^ that compiled predictors of severe COVID-19 outcome. For each patient, we queried the medication history table to identify use of any antidepressant, and we used the concept relation tables to match antidepressants to their active ingredients, resulting in 18 antidepressants that each had more than 1000 users. For each user we obtained 1) history of factors that might influence severe COVID-19 outcome, collected before positive COVID-19 test, denoted as the set of variables *L*; 2) which antidepressants they used before or at time of COVID-19 diagnosis; 3) presence of severe COVID-19 outcome (defined as severe hospitalization including ventilation, or death), as compared to non-severe COVID-19 outcome (no hospitalization, or hospitalized without ventilation).

Next, we identified cohorts for each of our 153 emulated trials. Each trial compared COVID-19 positive people taking one of two antidepressants. Users taking either of the two drugs were assigned to one of the two arms, and those taking both drugs were removed from the study. We assessed the imbalance between the two treatment groups within a cohort, for each risk factor *L* in *L*_*i*_ as the standardized absolute difference in frequency: 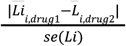. We also calculate the difference in the frequency of severe outcome between the two cohorts.

### Prediction of counterfactual outcomes and average treatment effects

The set of cohort data including *L*, treatment assignment *A*, and severity as a binary outcome was used to train a range of models for counterfactual inference.factor data was used as input to the counterfactual inference models. These models yield a counterfactual outcome, also known as potential outcome, for each individual, estimating their probability of disease if they had taken either of the two drugs. For each model, we performed hyperparameter tuning using cross-validation and obtain the best fit model for the next stage. Next, we use a held out portion of the data to predict counterfactual outcomes for treatment effect estimation. The difference of these counterfactual outcomes comprises the individual treatment effect. We average the individual treatment effects to obtain an average treatment effect. The confidence intervals are obtained by resampling the held out set with replacement and obtaining the average treatment effects in these bootstrap samples.

### Simulation of collider bias in counterfactual inference methods

We hypothesize that the association between ITE and number of measured risk factors could be due to a collider bias. To demonstrate this hypothesis, we simulated a data set containing 10 randomly assigned risk factors, each with p=0.2 of occurring in a given patient. If each risk factor independently impacts both drug assignment and outcome, then we can simulate the probability of both of these with an additive model. In order to create a simulation where the mean probability of drug exposure and outcome was 0.5 we used the following formula: 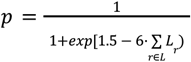. Given this value for *p* for an individual, we then simulated both exposure and outcome by sampling from a Bernoulli with the appropriate value of *p*.

We simulated data sets with 10,000 individuals. Then, we hid varying numbers of the 10 risk factors and performed our counterfactual effect estimation as above.

## Results

### Antidepressant-wide association studies of COVID-19 severe outcome

We extracted people diagnosed with COVID-19 in the N3C cohort, and among this population we identified 18 common antidepressants. In order to assess their association with severe COVID-19 outcome, we conducted a systematic set of target trial emulations, each emulating a clinical trial to test the effect of an antidepressant as compared to an active comparator, for a total of 153 trials. We chose to emulate an active comparator analysis as this identifies eligible participants who should be somewhat similar to each other, as they all are likely to be treated for some mental health condition. Our high-throughput target trial emulation approach is described in Table 1. Across 153 cohort studies, the median number of individuals participating was 104145, with a median of 9276 taking the less common drug (Figure 1A).

**Table 1:**
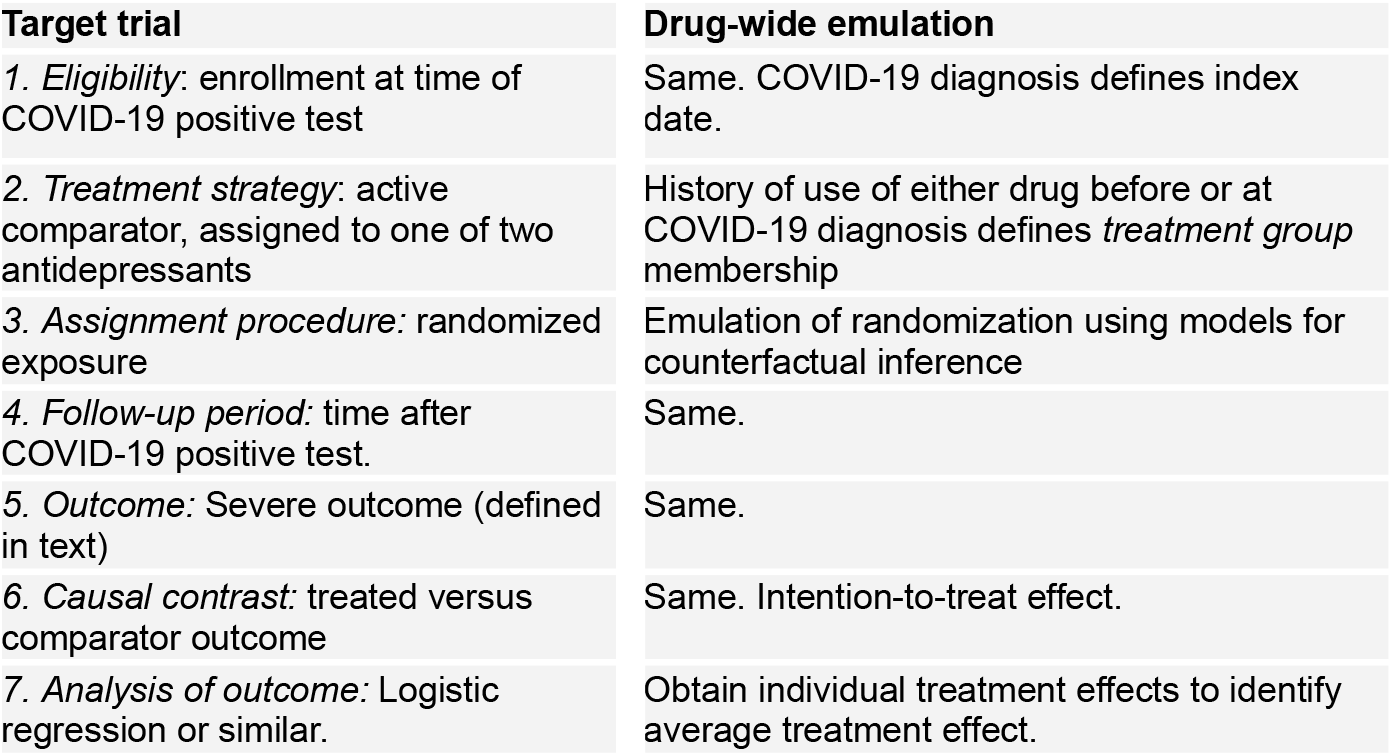
Procedure for emulating the seven components of the target trial.

**Figure 1.**
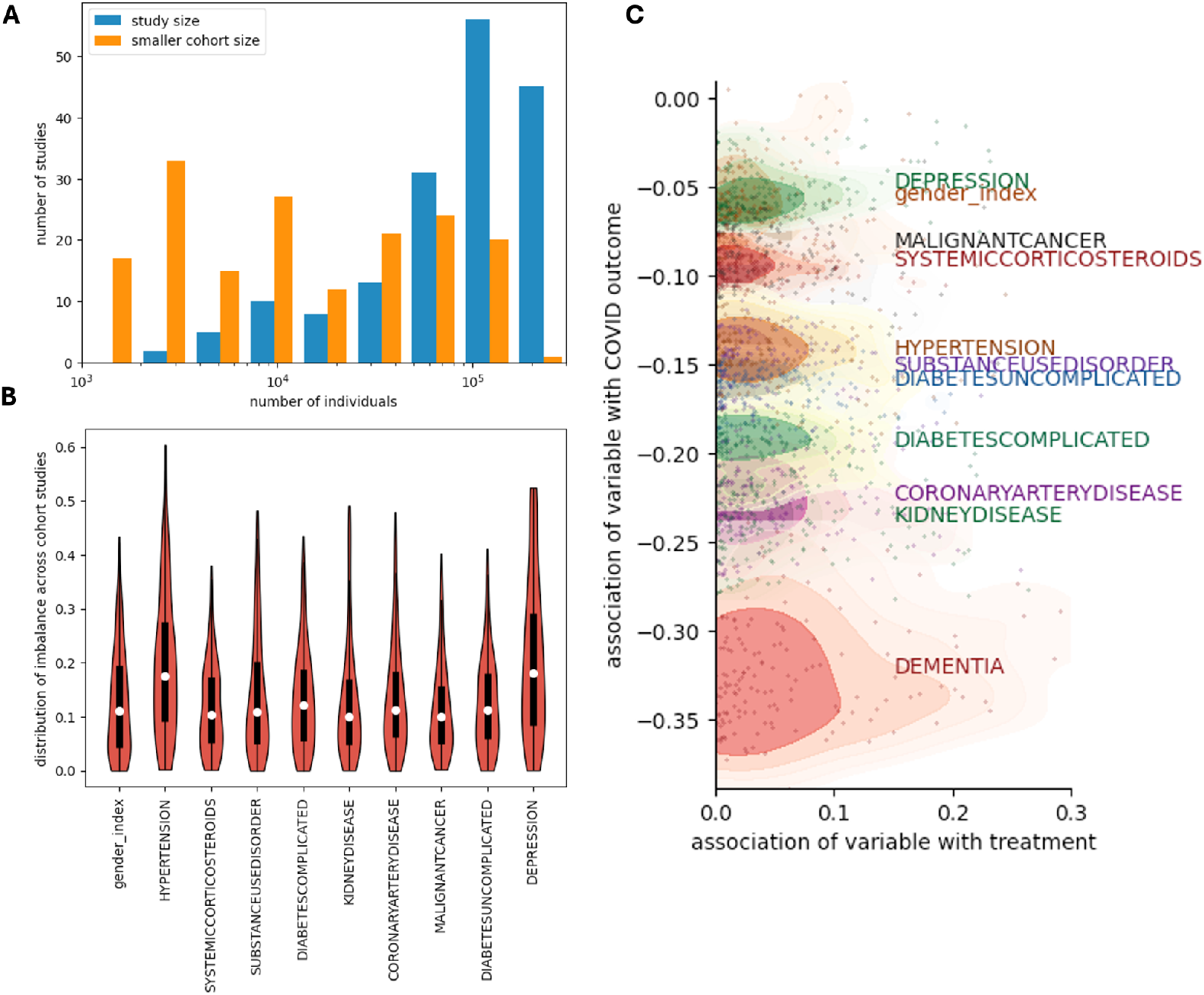
Overview of anti-depressant wide association studies and risk of confounding bias. **A**. Overview of the cohort sizes for each of the 153 active comparator studies. Each study compares two antidepressant cohorts, so for each study the cohort size of the smaller group is also shown. **B**. We compiled the risk factors for severe outcome that were also associated with imbalance between the two treatment groups. The factors shown had a median standardized absolute difference > 0.05. **C**. For each combination of a risk factor and a cohort study we assessed 1) absolute difference in frequency of the risk factor between the two groups (similar to B, x-axis), and 2) undadjusted association of the risk factor with severe outcome. Then, each point represents the potential for each risk factor to confound each cohort study (colored by risk factor). Dementia has by far the strongest association with severe outcome.

We first assessed imbalance for risk factors for severe outcome between the two cohorts, for each of the cohort studies. Out of 32 risk factors considered, we found that 10 had imbalance in at least half of the cohort studies, using an imbalance cutoff of greater than 5% standardized difference between the two cohorts (Figure 1B). Next, we assess for each risk factors, its potential to induce confounding. Confounding would be expected to occur when a risk factor is associated with both drug exposure and with outcome. Therefore, for each cohort, we assess for each risk factor its association with drug treatment assignment (x-axis, Figure 1C), and its association with COVID-19 outcome (y-axis, Figure 1C). We find that although dementia was not one of the most imbalanced risk factors across all studies, it has a high potential to induce confounding bias for some studies because it is so strongly associated with severe outcome.

### Application of a range of methods to estimate individual treatment effect in each cohort study

For each study, we apply a range of methods for counterfactual inference in order to estimate the individual treatment effect and, across the population, the average treatment effect. The methods have been surveyed elsewhere^9^, but we briefly summarize them. First, counterfactual estimation generally models the probability of outcome as a function of the supplied set of risk factors, both in the scenario where the individual was treated with the treatment, and in the case of treatment with the comparator. Then, the counterfactual outcome is predicted for each individual, in each of the two scenarios. If assumptions for causality are met, then the predicted counterfactual outcome should be independent of treatment assignment, conditional on the risk factors. The counterfactual outcomes are compared to obtain the individual treatment effect. These methods have been categorized in a framework describing “metalearners”^10^. In the T-learner, classification models are trained to predict outcome as a function of the risk factors in each of the two groups, using data from each group to fit a separate model. In the S-learner, only one model is trained, and treatment is included along with risk factors to predict outcome. Both T-learner and S-learner are agnostic as regards the classification model, and we experiment with including either logistic regression (T-LR, and S-LR) or random forest (T-RF and S-RF). A natural extension of these models is to use deep learning to perform the classificaiton task. TarNet^11^ uses multi-layer perceptrons to learn a latent space representing both the treatment groups before branching to learn separate classifiers for each of the two groups, somewhat analogous to the T-learner. As well, CEVAE, the causal effect variational autoencoder model, replaces the multi-layer perceptron with a variational autoencoder, a model that also explicitly seeks latent variables representing the true factors influencing both treatment and outcome.

We apply our target trial emulation approach (Table 1) using each of these six models in step 3, meaning they are all applied to identical sets of cohort studies. We obtain the individual treatment effects, average treatment effects, and bootstrap based confidence intervals (Methods). In order to compare the methods and assess if the risk factors confound the association of antidepressants and severe outcome, for each study we identify 1) the unadjusted absolute association of treatment exposure and outcome (Fig 2A, x-axis), 2) the strongest association of any confounding risk factor with exposure (Fig 2A, y-axis). For each target trial, we also plot the estimated average treatment effect (ATE), in Figure 2A, for the T-LR model (Fig 2A, color). We observe a clear association where studies with the greatest risk of confounding (highest values on x- and y-axes) also have the strongest estimated ATE. As dementia appeared as a particularly strong potential confounder, we also repeated the calculations for Figure 2A but used dementia history, rather than the strongest confounder for any study. The result (Figure 2B) shows that dementia is strongly associated with exposure in a subset of cohort studies, and these studies all have the strongest estimated ATE in the T-LR model. All of these studies compare mirtazapine against another antidepressant. In Figure 2C,D, we show that all of the counterfactual estimation methods share this pattern.

**Figure 2:**
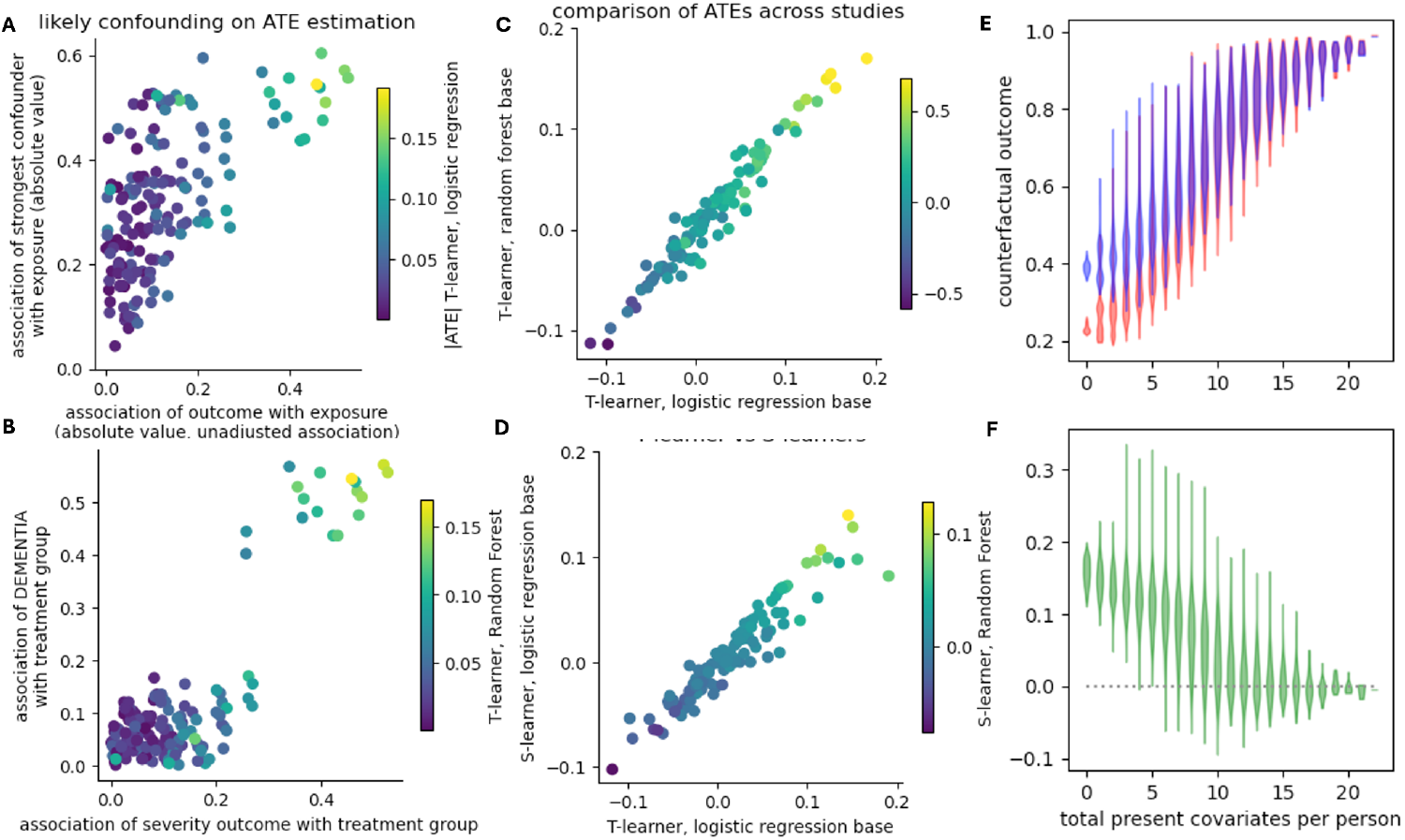
Evidence of confounded ATE. **A**. For each target trial (points) we estimate the association of the outcome with exposure in terms of absolute difference (x-axis), and we obtain the strongest association of any risk factor with exposure (y-axis), as well as the ATE obtained from T-LR for that trial. Emulated trials with the strongest chance of suffering confounding (high association of a risk factor with treatment, y-axis) also have the highest unadjusted association (x-axis) and the highest ATE (color scale). **B**. The effect is even more pronounced when only dementia as a confounder is considered. The studies where dementia is associated with treatment group all have the highest ATE, and these studies all compare mirtazapine against another antidepressant. **C, D**. ATEs are largely consistent across our five methods. **E**,**F**. Individuals with the fewest measured risk factors have the strongest ITE.

Because our previous analyses led us to strongly suspect confounding, we investigated the source of the strong ATEs. While we suspected that improper ITE estimation among dementia patients might be to blame we were surprised to find that the ITE was not associated with dementia or any other of the risk factors. However, we did observe that ITE was strongly associated with the total number of risk factors observed in a patient (Figure 2E, F). That is, the more risk factors observed (Figure 2E,F x-axis), the closer the ITE was to the null (Figure 2F, y-axis).

### Simulation suggests that conditioning on treatment exposure creates a collider bias due to unmeasured confounding

Investigating explanations for the association of ITE with number of observed risk factors, we sought to understand for the puzzling signal estimating that people with no observed risk factors have the strongest drug effect. As logistic regression is a linear model, the result suggested that the intercept was somehow inflated or deflated between the two exposure groups. Therefore, we asked whether the fact that models for counterfactual inferences are somehow conditioning on treatment exposure could induce a bias in some cases. That is, when many factors impact both exposure and outcome, conditioning on observed risk factors creates a back door path (Figure 3A). To test this idea, we simulated a data set containing 10 risk factors, and hid three of them from the model training. We recovered both the expected inflated ATE and the expected negative association between observed and unobserved number of risk, conditioned on treatment assignment. Furthermore, our simulation results mirror the findings of our study: peopel with fewer observed risk factors have higher ITE, particularly in the logistic regression models but also in the random forest model (Figure 3B). This suggests that poorly modeled unmeasured confounding can create this pattern. To our knowledge, this is the first time this has been reported, and it can serve as a useful signal of poorly controlled confounding.

**Figure 3:**
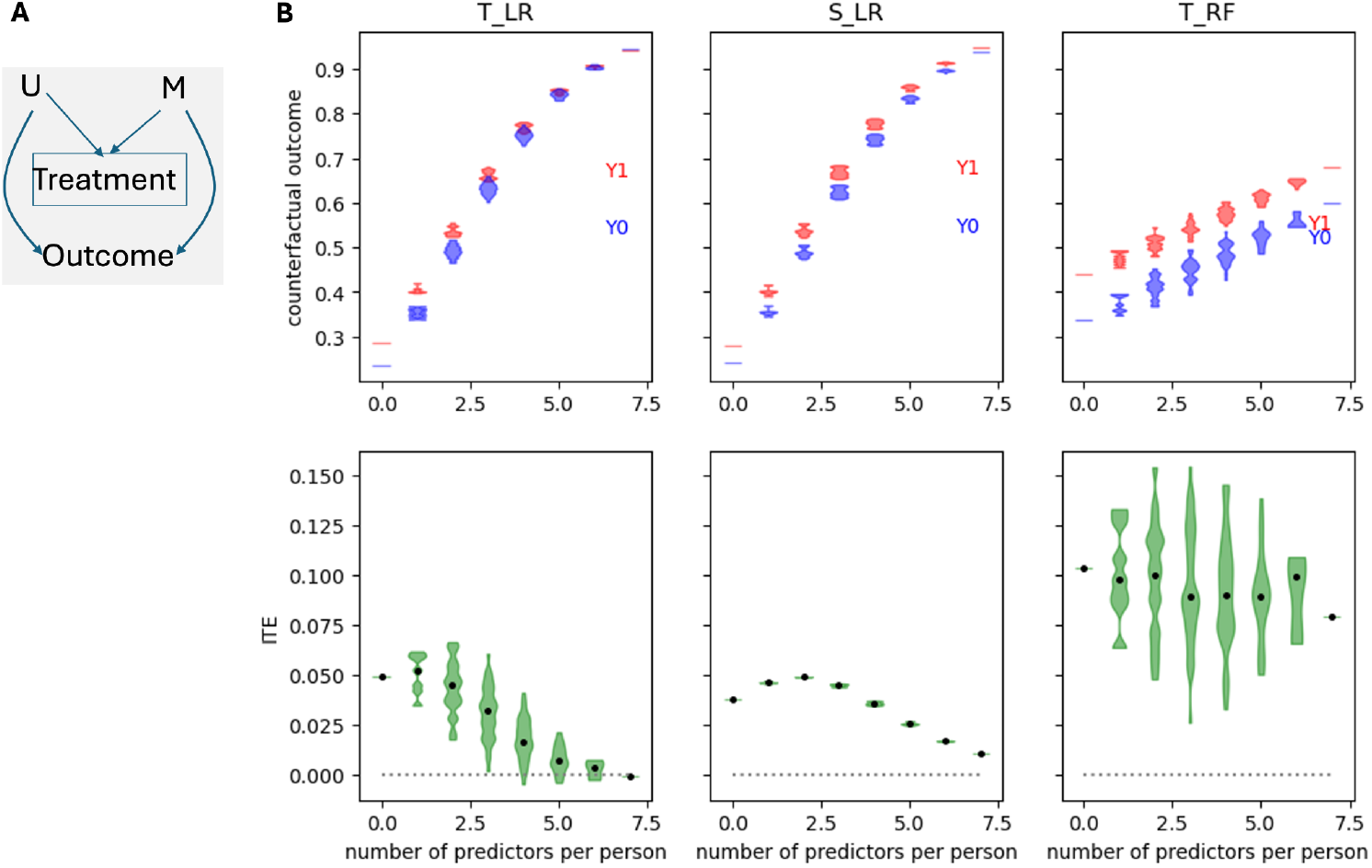
Simulation and theory behind collider bias inflating ITE and ATE. **A**. Conditional on treatment (outlined box), unmeasured confounders (U) are more common in those without measured confounders (M), creating a biased association between treatment and outcome. **B**. Simulating binary risk factors, treatment, and outcome following this model, and hiding three of the risk factors from the model results in inflated ATE and an association between number of measured risk factors (x-axis) and ITE (y-axis), bottom, for all three models although particularly for logistic regression.

### Antidepressant-wide study finds no convincing association with COVID-19 outcome

All of the effects with signficant bootstrap interval were comparator studies of mirtazapine, suggesting that if our study is to point to any effects on COVID-19 outcome it would be among mirtazapine users. We focused our analysis to see if any consistent effect with mirtazapine could be identified. Across all 17 active comparator trials of mirtazapine, we observed an elevated ATE across all estimators except for the S-learner with logistic regression base. While the bootstrap intervals did not overlap the null for the metalearners, for Tarnet the ATE was much higher, but also the confidence intervals were much wider (Figure 4, bottom). This suggests that effect estimation with Tarnet is less stable.

**Figure 4:**
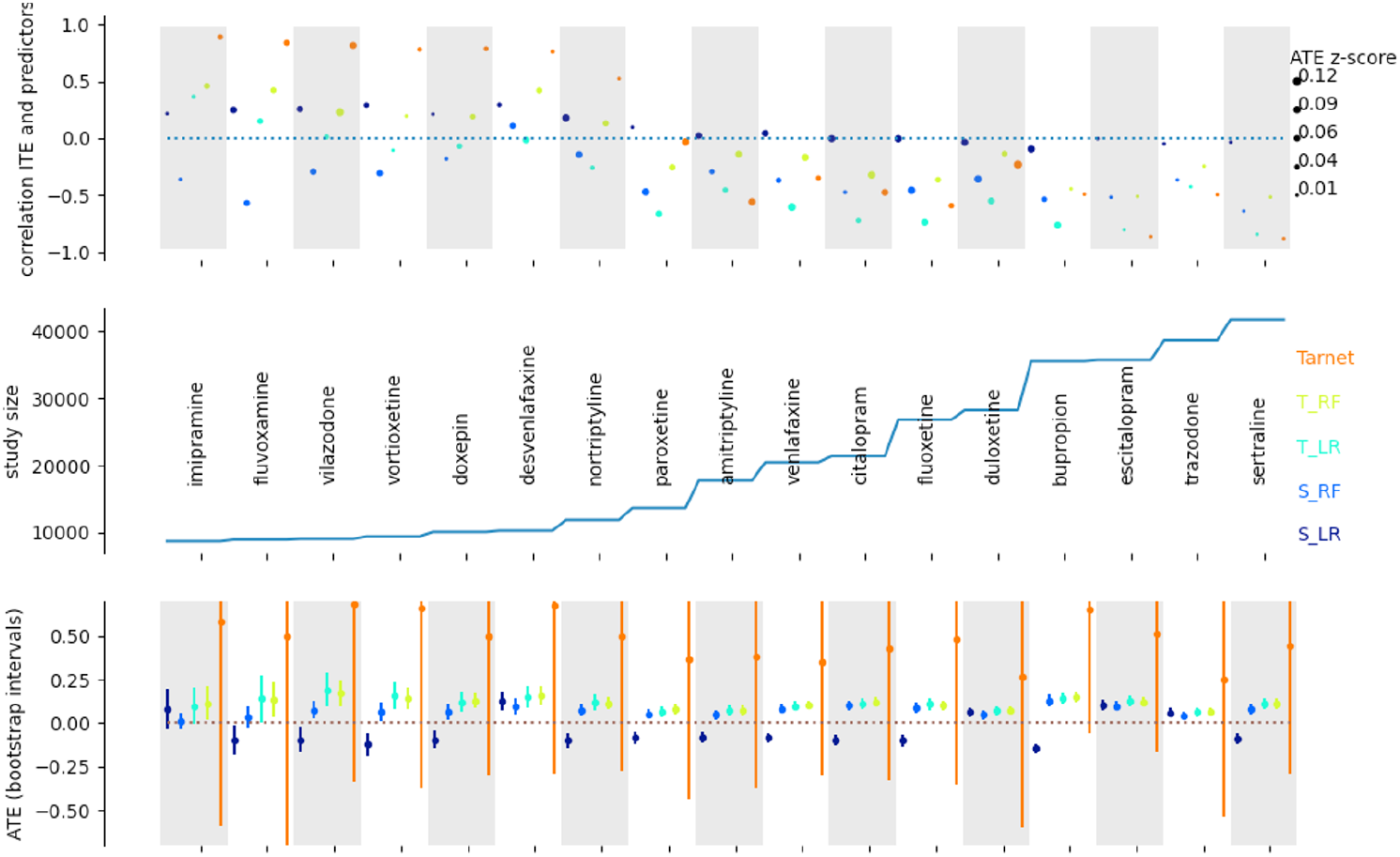
Investigating the results of emulating active comparator target trials of mirtazapine.

We assessed whether the correlation of ITE with total number of measured risk factors was present across all of these cohort studies (Figure 4, top). We found that in general, across most methods, there was a strong negative correlation of ITE with total number of risk factors, similar to our result in Figure 2F. This suggests that all of the mirtazapine comparisons may be subject to the same collider bias. We are not able to confidently identify any antidepressant associated with COVID-19 outcome.

## Discussion

We have conducted, to our knowledge, the only study systematically comparing the results of multiple methods for counterfactual inference in real-world data. Our results show that the results of these methods are highly correlated, but also that they have distinctive properties and potential for bias. By applying multiple methods, we are able to assess the consistency between them. Additionally, by emulating a large number of target trials simultaneously, we discover patterns suggesting unmeasured confounding. This supports the potential of large-scale target trial emulation for assessing and improving methods for causal effect discovery.

Our approach does have some weaknesses. First, we did not attempt to adjust for all confounders that could influence medication use. However, our study did include almost all variables adjusted for in a recent study of SSRIs and COVID-19^5^. Because we sought to systematically perform the analysis for all antidepressants, we chose to include the set of variables that have been determined to impact COVID-19 outcome, and we did find a strong effect of adjusting for these variables on the association between severe outcome and treatment. Second, and relatedly, drug-wide studies cannot tailor the target trial emulation to each comparison (in our case, 153 comparisons). Although this weakness means that we may not include some variable that an expert would adjust for in the target trial emulation, we believe that the systematic and objective nature of our analysis is a strength. Drug-wide analyses can always be followed by tailored expert designed analysis, but drug-wide analysis is useful both for discovering unexpected effects, and for providing a basis for comparing methods and identifying their potential weaknesses.

This approach has potential to guide analyses emulating target trials using counterfacutal inference, and other methods. Our finding that the overall burden of measured variables is associated with ITE can serve as a useful signal of unmeasured confounding for other studies. We have contributed a structural explanation for this signal of bias, based on the collider structure that results from conditioning on observed treatment assignment. This signal can inform evaluation of these methods and contribute to improved designs of counterfactual inference approaches.

Finally, we assessed the association of antidepressant use with COVID-19 outcomes using an active comparator design. By performing an antidepressant-wide association analysis we were able to make a number of conclusions.

First, antidepressants vary with respect to their association with risk factors for severe outcome. A number of the risk factors were associated with one or more antidepressants. Particularly mirtazapine is associated with history of dementia use, meaning the cohort of mirtazapine users is particularly at risk for severe outcome.

Second, while counterfactual inference methods do indeed adjust well for measured confounders such as dementia, unmeasured confounders can still impact the estimates of average treatment effect, and it appears that collider bias induced by conditioning on treatment may play a role in these biased estimates. This observation alone can serve as a valuable contribution, as there are few methods for evaluating the results of observational studies, as true drug effects can only be known via clinical trials that have not yet been performed. We hypothesize that the true issue is not the conditioning on treatment effect, but that the methods are not adequately modeling bias due to unmeasured confounding. It is possible that if we had included a wider range of predictors in the models, then we could have created estimates more resistant to bias. This could be a topic for future work, which could be easily compared to our results.

Third, drug (antidepressant)-wide association studies are a valuable approach for assessing bias and making robustly supported conclusions. Because we systematically compared all pairs of antidepressants, we were able to discover this pattern. While one method evaluates drug effects by comparing them to *negative controls* or performing outcome-wide analysis and assuming all effects are subject to the same bias^12^, our approach instead uncovers bias in the drug effects most associated with outcome. Although drug-wide analyses do not allow for expert design that aims to guarantee a causal interpretation of drug-outcome associations, we have shown they can provide valuable insight.

Fourth, we conclude that we are not able to identify any meaningful effect of any antidepressant on COVID-19 outcome. While mirtazapine appears consistently associated with poor outcome across all comparator drugs, our analyses suggest that this is most likely due to unmeasured confounding. It seems likely that as this drug appears to be given to elderly people who were known to be particularly vulnerable to the effects of COVID-19, they may have comorbidities that were not measured or included in the data set.

Overall, we have presented a rigorous analysis of the possible effects of antidepressants on COVID-19 outcome, we have performed systematic application of a range of methods for counterfactual inference in a large scale target trial emulation study, and we have developed a new approach for comparing these methods to assess their weaknesses and assess support for drug effects. These findings should guide future work emulating target trials using real-world data.

## Data Availability

All data analyzed are available online at https://covid.cd2h.org/enclave/
No data were generated by this study.

## Notes

### Competing Interest Statement

The authors have declared no competing interest.

### Funding Statement

This work was supported by the National Institute of General Medicine Sciences (NIGMS R35 GM151001-01) to RDM.

### Author Declarations

The study used ONLY openly available human data that were originally located at: https://covid.cd2h.org/enclave/

